# COVID-19 vaccination effectiveness rates by week and sources of bias

**DOI:** 10.1101/2021.12.22.21268253

**Authors:** Anna Ostropolets, George Hripcsak

## Abstract

**Importance:** Randomized clinical trials and observational studies have demonstrated high overall effectiveness for the three US-authorized COVID-19 vaccines against symptomatic COVID-19 infection. Nevertheless, the challenges associated with the use of observational data can undermine the results of the studies.

**Objective:** To assess the feasibility of using observational data for vaccine effectiveness studies by examining granular weekly effectiveness.

**Design, Settings and Participants:** In this retrospective cohort study, we used Columbia University Medical Center data linked to State and City Immunization Registries to assess the weekly effectiveness of mRNA COVID-19 vaccines. We conducted manual chart review of cases in week one in both groups along with a set of sensitivity analyses for Pfizer-BioNTech, Moderna and Janssen vaccines.

**Main Outcomes and Measures:** We used propensity score matching with up to 54,987 covariates and fitted Cox proportional hazards models to estimate hazard ratios and constructed Kaplan-Meier plots for two main outcomes (COVID-19 infection and COVID-19-associated hospitalization).

**Results:** The study included 179,666 patients. We observed increasing effectiveness after the first dose of mRNA vaccines with week 6 effectiveness approximating 84% (95% CI 72-91%) for COVID-19 infection and 86% (95% CI 69-95) for COVID-19-associated hospitalization. When analyzing unexpectedly high effectiveness in week one, chart review revealed that vaccinated patients are less likely to seek care after vaccination and are more likely to be diagnosed with COVID-19 during the encounters for other conditions. Sensitivity analyses showed potential outcome misclassification for COVID-19 ICD10-CM diagnosis and the influence of excluding patients with prior COVID-19 infection and anchoring in the unexposed group. Overall vaccine effectiveness analysis in fully vaccinated patients matched the results of the randomized trials.

**Conclusions and Relevance:** Observational data can be used to ascertain vaccine effectiveness if potential biases are accounted for. The data need to be scrutinized to ensure that compared groups exhibit similar health seeking behavior and are equally likely to be captured in the data. Given the difference in temporal trends of vaccine exposure and baseline characteristics, indirect comparison of vaccines may produce biased results.

**KEY POINTS:** *Question:* When accounted for all potential biases, what is the weekly effectiveness of COVID-19 vaccines?

*Findings:* In this cohort study we replicated the results of randomized clinical trials, discovered plausible increase in effectiveness after week one following the first dose of mRNA COVID-19 vaccines and found differences in temporal trends of vaccine exposure and baseline characteristics in vaccinated groups.

*Meaning:* Observational data can be used to reliably estimate vaccine effectiveness if the biases are accounted for. Vaccines need to be directly compared.

## BACKGROUND

Randomized clinical phase-3 trials have demonstrated high efficacy for the three US-authorized COVID-19 vaccines against symptomatic COVID-19 infection, ranging from 66.9% for Ad26.COV2.S (Johnson & Johnson–Janssen) to 94.1% and 94.6% for BNT162b2 (Pfizer–BioNTech) and mRNA-1273 (Moderna) vaccines ^1–3^. Their fast approval and widespread use require robust post-marketing studies that leverage large sample size, heterogeneous populations, and longer follow-up available in observational data.

There have been several recent observational studies, which have shown effectiveness similar to the randomized clinical trials (RCTs). Thompson et al. used a test-negative design to examine the effectiveness of Pfizer–BioNTech and Moderna vaccines with respect to COVID-19 hospitalization across a network of institutions ^4^. The cohort study by Tartof et al. examined the effectiveness of Pfizer– BioNTech against COVID-19 infection and hospitalization in fully vaccinated patients, reporting the limitations of matching the vaccinated and unvaccinated populations ^5^. Another cohort study by Polinski et al. used a large population to assess the effectiveness of Ad26.COV2.S and obtained similar results despite the fact that the data source did not allow to ascertain vaccination status for all patients ^6^. There were several non-US studies showing similar overall effectiveness, which nevertheless may not be generalizable to the US population due to differences in patient populations, COVID-19 variants spread and baseline COVID-19 prevalence ^7,8(p2),9–11^.

While the existing observational studies matched randomized clinical trial results, there is a growing number of pressing questions related to COVID-19 vaccine effectiveness such as effectiveness against new variants and vaccine durability, for which trials may not be readily available ^12^. Moreover, the challenges associated with the use of observational data such as incomplete data capture, outcome misclassification and appropriate comparator sampling can undermine the results of the studies if such biases are not accounted for ^13(p)^. An example of such a challenge is the estimation of vaccine effectiveness during the first two weeks following the first dose. Studies have shown contradicting results for Pfizer–BioNTech vaccine with effectiveness ranging from moderate effectiveness of 52% ^3^ to very high effectiveness of 92.6% ^14^.

The goal of this study was to assess the feasibility of using observational data for vaccine effectiveness studies by examining more granular weekly effectiveness estimates and uncovering underlying biases and challenges. We employed large-scale propensity score matching and many negative controls to reduce and assess bias, and we leveraged a range of sensitivity analyses as well as manual review of the COVID-19 infection cases during the first week after vaccination.

## METHODS

### Main design

For this retrospective observational cohort study, we used electronic health records from the Columbia University Irving Medical Center (CUIMC) database (Appendix 1), which has an ongoing automated connection to New York City and State public health department vaccine registries and includes all within-state vaccinations for our population. The data were translated to the OMOP Common Data Model version 5 and was previously used in multiple studies ^15^.

We studied the three main US COVID-19 vaccines separately. Three target cohorts included patients indexed on the first dose of one of the corresponding vaccines with no prior COVID-19 infection and no previous exposure to other COVID-19 vaccines. Our comparator group was unvaccinated patients who were indexed on a date selected from the unvaccinated patient’s history (not necessarily with any medical event) such that it matched the index date of one of the target group participants. Both the target and comparator groups had at least 365 days of prior observation and primarily resided in New York.

Outcomes of interest included a) COVID-19 infection defined as a positive COVID-19 test (e.g., reverse-transcriptase–polymerase-chain-reaction assays) or a diagnostic code of COVID-19 and b) COVID-19 hospitalization defined as an inpatient visit associated with a COVID-19 positive test or diagnosis within 30 days prior or during the visit. Upon further examination of the results, we added two other outcomes: a) COVID-19 positive test only and b) COVID-19 hospitalization associated with a positive COVID-19 test. Design overview is provided in Appendix 2; code lists and links to phenotype definitions are provided in Appendix 3.

For the time-at-risk, we selected six consecutive 7-day intervals after the first dose until an outcome, end of observation period or death, whichever came earlier. Additionally, given the results for vaccine effectiveness during week 1 following the first dose, we conducted chart review for patients with a COVID-19 positive test recorded in the abovementioned period. We reviewed all cases for the vaccinated population as well a random sample of the cases in the unvaccinated population.

### Sensitivity analyses

Along with studying granular weekly intervals, we assessed overall absolute vaccine effectiveness in patients with at least one dose of a COVID-19 vaccine and in fully vaccinated patients. The latter was defined as 14 days after the second dose of Pfizer-BioNTech or Moderna vaccines or first dose of Janssen vaccine. For each comparison we estimated hazard ratios (HRs) and constructed Kaplan-Meier plots as described below.

Given that the published studies focused on patients without prior COVID-19 infection, our second sensitivity analysis included all eligible patients regardless of their previous COVID-19 status. Finally, as the strategy for unvaccinated group index date selection (anchoring) has been reported to influence incidence of outcomes ^16^, we additionally tested an unvaccinated comparator indexed on a healthcare encounter matching the index date of one of the target group participants within 3 days corridor, with at least 365 days of prior observation located at New York.

### Statistical methods

For each analysis, we fitted a lasso regression model to calculate propensity score and match patients in each target and comparator group with 1:1 ratio. For propensity model we used all demographic information, index year and month, as well as the number of visits, condition and drug groups, procedures, device exposures, laboratory and instrumental tests and other observations over long (prior year) and short-term period (prior month).

For each outcome, we fitted a Cox proportional hazards models to estimate HRs and constructed Kaplan-Meier plots. Empirical calibration based on the negative control outcomes was used to identify and minimize any potential residual confounding by calibrating HRs and 95% confidence intervals (CIs) ^17,18^. Vaccine effectiveness was calculated as 100% × (1−hazard ratio).

All analyses were supported by the OHDSI Infrastructure (CohortMethod package, available at https://ohdsi.github.io/CohortMethod/, FeatureExtraction available at https://ohdsi.github.io/FeatureExtraction/ and the Cyclops package for large-scale regularized regression ^19^ available at https://ohdsi.github.io/Cyclops).

### Diagnostics

We used multiple sources of diagnostics to estimate potential bias and confounding following best practices for evidence generation ^20^. First, we examined covariate and propensity score balance prior to proceeding with outcome modelling and effect estimation to ensure that we have enough sample size and to control for potential observed confounding ^20^. We plotted propensity scores to investigate the overlap in patient populations at the baseline and examined the balance of all baseline characteristics to determine if the target and comparator cohorts were imbalanced at the baseline and after propensity score matching. Target and comparator cohorts were said to be balanced if the standardized difference of means of all covariates after propensity score matching was less than 0.1 ^21^.

For negative control calibration, we used 93 negative controls (Appendix 4) with no known causal relationship with the COVID-19 vaccines. Negative controls were selected based on a review of existing literature, product labels and spontaneous reports and were reviewed by clinicians ^22^. We assessed residual bias from the negative control estimates.

## RESULTS

### Patient characteristics

In total, we identified 179,666 patients with at least one dose of COVID-19 vaccine: 121,771 patients for Pfizer-BioNTech, 52,728 for Moderna and 5,167 for Janssen (Table 1).

**Table 1.** Patient baseline characteristics for patients with at least one dose of a COVID-19 vaccine and the comparator unvaccinated patients, before and after propensity score matching.

|  | Before matching |  |  |  |  |  |  |  |  | After matching |  |  |  |  |  |  |  |  |
| --- | --- | --- | --- | --- | --- | --- | --- | --- | --- | --- | --- | --- | --- | --- | --- | --- | --- | --- |
|  | Pfizer |  |  | Moderna |  |  | Janssen |  |  | Pfizer |  |  | Moderna |  |  | Janssen |  |  |
| Characteristic | Ta<br>rge<br>t | Com<br>parat<br>or | Std<br>.diff | Ta<br>rge<br>t | Com<br>parat<br>or | Std<br>.diff | Ta<br>rge<br>t | Com<br>parat<br>or | Std<br>.diff | Ta<br>rge<br>t | Com<br>parat<br>or | Std<br>.diff | Ta<br>rge<br>t | Com<br>parat<br>or | Std<br>.diff | Ta<br>rge<br>t | Com<br>parat<br>or | Std<br>.diff |
| Patients, n | 12<br>1,7<br>71 | 164,9<br>97 |  | 52,<br>72<br>8 | 148,7<br>95 |  | 5,1<br>67 | 52,64<br>3 |  | 10<br>1,1<br>09 | 101,1<br>11 |  | 50,<br>51<br>7 | 50,51<br>7 |  | 5,0<br>31 | 5,031 |  |
| Follow-up, days.<br>Median (IQR) | 10<br>7<br>(80<br>–<br>13<br>7) | 104<br>(71-137) |  | 12<br>7<br>(1<br>02<br>–<br>15<br>3) | 123<br>(99-153) |  | 79<br>(7<br>2-95<br>) | 79<br>(72-95) |  | 10<br>7<br>(78<br>–<br>14<br>9) | 107<br>(79-140) |  | 12<br>6<br>(1<br>01<br>–<br>15<br>3) | 126<br>(102-153) |  | 79<br>(7<br>2-95<br>) | 79<br>(72-95) |  |
| *Age group, %* |  |  |  |  |  |  |  |  |  |  |  |  |  |  |  |  |  |  |
| 10-19 | 4.2 | 10.8 | -<br>0.2<br>5 | 0.5 | 1.7 | -<br>0.1<br>2 | 0.8 | 0.8 | 0.0<br>0 | 4.8 | 4.3 | 0.0<br>2 | 0.5 | 0.4 | 0.0<br>1 | 0.8 | 0.8 | 0.0<br>0 |
| 20-49 | 37.<br>2 | 42.6 | -<br>0.1<br>1 | 35.<br>7 | 45.7 | -<br>0.2<br>0 | 43.<br>9 | 43 | 0.0<br>2 | 40.<br>3 | 40.1 | 0 | 36.<br>9 | 37.4 | -<br>0.0<br>1 | 44.<br>2 | 43.9 | 0.0<br>1 |
| 50-64 | 23.<br>9 | 20.3 | 0.0<br>9 | 21.<br>2 | 23.3 | -<br>0.0<br>5 | 31.<br>7 | 31.7 | 0.0<br>0 | 23.<br>6 | 23.7 | 0 | 21.<br>7 | 21.4 | 0.0<br>1 | 31.<br>8 | 31.3 | 0.0<br>1 |
| 65-74 | 18.<br>8 | 12.6 | 0.1<br>7 | 21.<br>3 | 14.4 | 0.1<br>8 | 11.<br>6 | 12.2 | -<br>0.0<br>2 | 15.<br>8 | 16.6 | -<br>0.0<br>2 | 20.<br>6 | 20.5 | 0.0<br>0 | 11.<br>5 | 12 | -<br>0.0<br>2 |
| 75-84 | 11.<br>3 | 8.9 | 0.0<br>8 | 15.<br>4 | 10 | 0.1<br>6 | 7.6 | 7.9 | -<br>0.0<br>1 | 10.<br>6 | 10.7 | 0 | 14.<br>6 | 14.6 | 0.0<br>0 | 7.2 | 7.9 | -<br>0.0<br>3 |
| >84 | 4.1 | 3.8 | 0.0<br>2 | 5.8 | 4.8 | 0.0<br>4 | 4.3 | 4.3 | 0.0<br>0 | 4.2 | 4.1 | 0.0<br>1 | 5.6 | 5.6 | 0.0<br>0 | 4.2 | 4 | 0.0<br>1 |
| **Gender, %** |  |  |  |  |  |  |  |  |  |  |  |  |  |  |  |  |  |  |
| Female | 63.<br>7 | 57.8 | 0.1<br>2 | 64.<br>4 | 58.7 | 0.1<br>2 | 63.<br>4 | 63.2 | 0.0<br>1 | 61.<br>4 | 62 | -<br>0.0<br>1 | 64.<br>2 | 64.7 | -<br>0.0<br>1 | 63.<br>5 | 61.1 | 0.0<br>5 |
| **Race, %** |  |  |  |  |  |  |  |  |  |  |  |  |  |  |  |  |  |  |
| race = Asian | 3.8 | 2.6 | 0.0<br>7 | 4.2 | 2.8 | 0.0<br>7 | 3.6 | 1.7 | 0.1<br>2 | 3.5 | 3.4 | 0.0<br>1 | 4.2 | 4.4 | -<br>0.0<br>1 | 3.7 | 3.6 | 0.0<br>1 |
| race = Black or<br>African American | 12.<br>4 | 14.2 | -<br>0.0<br>5 | 8.7 | 14.2 | -<br>0.1<br>7 | 15.<br>9 | 15.5 | 0.0<br>1 | 12.<br>6 | 12.2 | 0.0<br>1 | 9 | 8.4 | 0.0<br>2 | 15.<br>7 | 15.5 | 0 |
| race = White | 40.<br>5 | 35.1 | 0.1<br>1 | 48.<br>3 | 34.4 | 0.2<br>9 | 37.<br>4 | 35.7 | 0.0<br>3 | 39.<br>3 | 39.5 | 0 | 46.<br>9 | 47.9 | -<br>0.0<br>2 | 37.<br>4 | 37.5 | 0 |
| **Medical history, %** |  |  |  |  |  |  |  |  |  |  |  |  |  |  |  |  |  |  |
| Chronic liver<br>disease | 0.6 | 0.6 | 0 | 0.5 | 0.6 | -<br>0.0<br>2 | 1.1 | 0.7 | 0.0<br>5 | 0.5 | 0.5 | 0 | 0.5 | 0.5 | 0 | 1 | 1.2 | -<br>0.0<br>2 |
| Chronic<br>obstructive lung<br>disease | 1.3 | 1 | 0.0<br>2 | 1.4 | 1.1 | 0.0<br>2 | 2.4 | 1.3 | 0.0<br>9 | 1 | 1 | 0.0<br>1 | 1.2 | 1.2 | 0 | 2 | 2.2 | -<br>0.0<br>1 |
| Dementia | 1.2 | 1.1 | 0 | 1 | 1.2 | -<br>0.0<br>2 | 2.6 | 1.1 | 0.1<br>1 | 1.1 | 1 | 0.0<br>1 | 1 | 0.9 | 0.0<br>1 | 2.2 | 2.2 | 0 |
| Depressive<br>disorder | 5.3 | 4 | 0.0<br>6 | 4.7 | 3.9 | 0.0<br>4 | 8 | 4.8 | 0.1<br>3 | 4 | 3.7 | 0.0<br>2 | 4.2 | 4 | 0.0<br>1 | 7.1 | 8 | -<br>0.0<br>3 |
| Diabetes mellitus | 7.1 | 5.2 | 0.0<br>8 | 6.6 | 5.6 | 0.0<br>4 | 10.<br>3 | 6.2 | 0.1<br>5 | 5.7 | 5.4 | 0.0<br>1 | 6.2 | 5.8 | 0.0<br>2 | 9.5 | 10.2 | -<br>0.0<br>2 |
| Human immunodeficiency virus infection | 1.4 | 1.1 | 0.0<br>3 | 0.9 | 1.2 | -<br>0.0<br>3 | 1.7 | 1.4 | 0.0<br>2 | 1.1 | 1 | 0 | 0.8 | 0.8 | 0 | 1.6 | 1.8 | -<br>0.0<br>1 |
| Hyperlipidemia | 12.<br>9 | 8.1 | 0.1<br>6 | 14.<br>9 | 8.9 | 0.1<br>9 | 14.<br>3 | 10.2 | 0.1<br>3 | 10.<br>2 | 9.5 | 0.0<br>2 | 13 | 12.6 | 0.0<br>1 | 13.<br>4 | 14.3 | -<br>0.0<br>3 |
| Hypertensive disorder | 16 | 11.3 | 0.1<br>4 | 16 | 12.4 | 0.1 | 21.<br>4 | 13.8 | 0.2 | 13.<br>1 | 12.2 | 0.0<br>3 | 14.<br>7 | 13.9 | 0.0<br>2 | 20.<br>1 | 21.7 | -<br>0.0<br>4 |
| Obesity | 5.1 | 4.9 | 0.0<br>1 | 4 | 4.4 | -<br>0.0<br>2 | 7.3 | 5.9 | 0.0<br>6 | 4.4 | 4.1 | 0.0<br>2 | 3.8 | 3.6 | 0.0<br>1 | 6.8 | 7.8 | -<br>0.0<br>4 |
| Osteoarthritis | 7.3 | 4.7 | 0.1<br>1 | 7.7 | 5.3 | 0.1 | 8.4 | 6.2 | 0.0<br>8 | 5.8 | 5.3 | 0.0<br>2 | 6.8 | 6.5 | 0.0<br>1 | 7.8 | 8.8 | -<br>0.0<br>4 |
| Renal impairment | 3.7 | 3 | 0.0<br>4 | 3.5 | 3.3 | 0.0<br>1 | 6.6 | 3.3 | 0.1<br>5 | 2.9 | 2.7 | 0.0<br>1 | 3.3 | 3 | 0.0<br>1 | 5.3 | 5.9 | -<br>0.0<br>2 |
| Cerebrovascular disease | 1.7 | 1.4 | 0.0<br>2 | 2.2 | 1.6 | 0.0<br>5 | 2.7 | 1.7 | 0.0<br>7 | 1.5 | 1.4 | 0.0<br>1 | 2 | 1.8 | 0.0<br>2 | 2.3 | 2.4 | -<br>0.0<br>1 |
| Heart disease | 8.6 | 7.1 | 0.0<br>6 | 10.<br>1 | 7.6 | 0.0<br>9 | 11.<br>8 | 8 | 0.1<br>3 | 7.5 | 7.1 | 0.0<br>2 | 9.2 | 8.7 | 0.0<br>2 | 10.<br>3 | 11.7 | -<br>0.0<br>4 |
| Malignant neoplastic disease | 5.3 | 4.5 | 0.0<br>4 | 6.5 | 5 | 0.0<br>7 | 5 | 4.9 | 0 | 4.7 | 4.3 | 0.0<br>2 | 5.9 | 5.5 | 0.0<br>2 | 4.8 | 5.2 | -<br>0.0<br>2 |

Among vaccinated patients, 68% received Pfizer-BioNTech vaccine, 29% received Moderna and 3% received Janssen vaccine. When investigating the vaccination pathways, we discovered that 112,963 patients (93% of patients with at least one dose of Pfizer-BioNTech) had 2 doses of Pfizer-BioNTech and 42,384 (80%) patients had 2 doses of Moderna. We found 344 and 291 patients with 3 doses of the corresponding vaccines and 440 patients having mixed Pfizer-BioNTech, Moderna and Janssen vaccines in different combinations.

Within our database, Moderna was administered early on with a peak in January 2021 (Figure 1), while Pfizer-BioNTech and Janssen vaccinations peaked in April. It was reflected in the follow-up time with Moderna patients having on average longer follow-up with some individuals having up to 5.8 months of post-observation.

**Figure 1.**
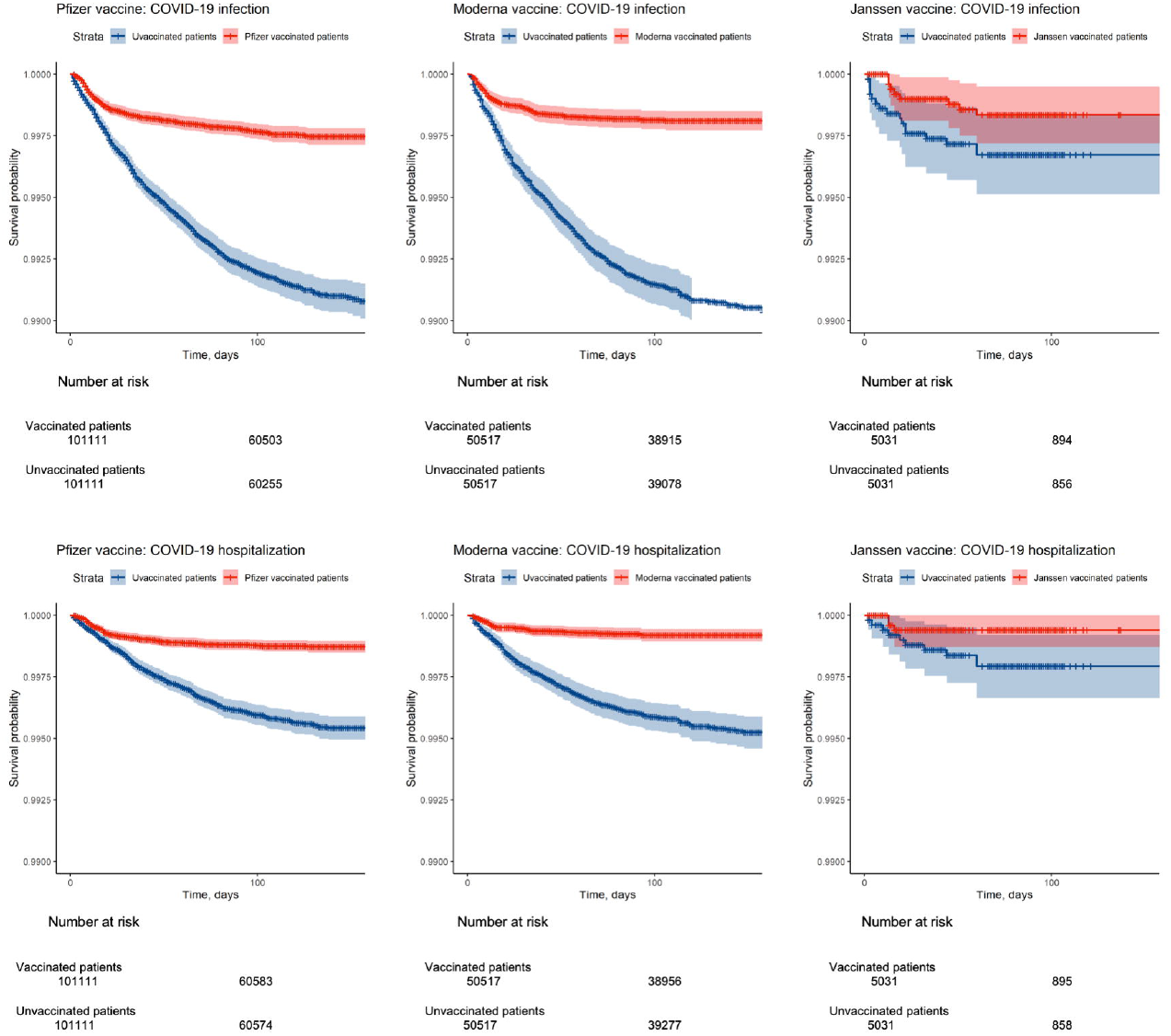
Distribution of vaccination month for COVID-19 vaccines. Black dots represent the number of incident COVID-19 cases (defined as a positive test) in each month.

We observed that unvaccinated comparator patients (Table 1) were on average younger and had fewer co-morbidities and less exposure to various drugs prior to matching. We were able to achieve balance on all covariates (up to 54,987 covariates, standardized difference of means less than 0.1) with propensity score matching. Figure 2 presents the covariate balance and propensity score balance plots showing that anchoring unvaccinated patients on a date allowed us to achieve better balance compared to anchoring patients on a visit.

**Figure 2.**
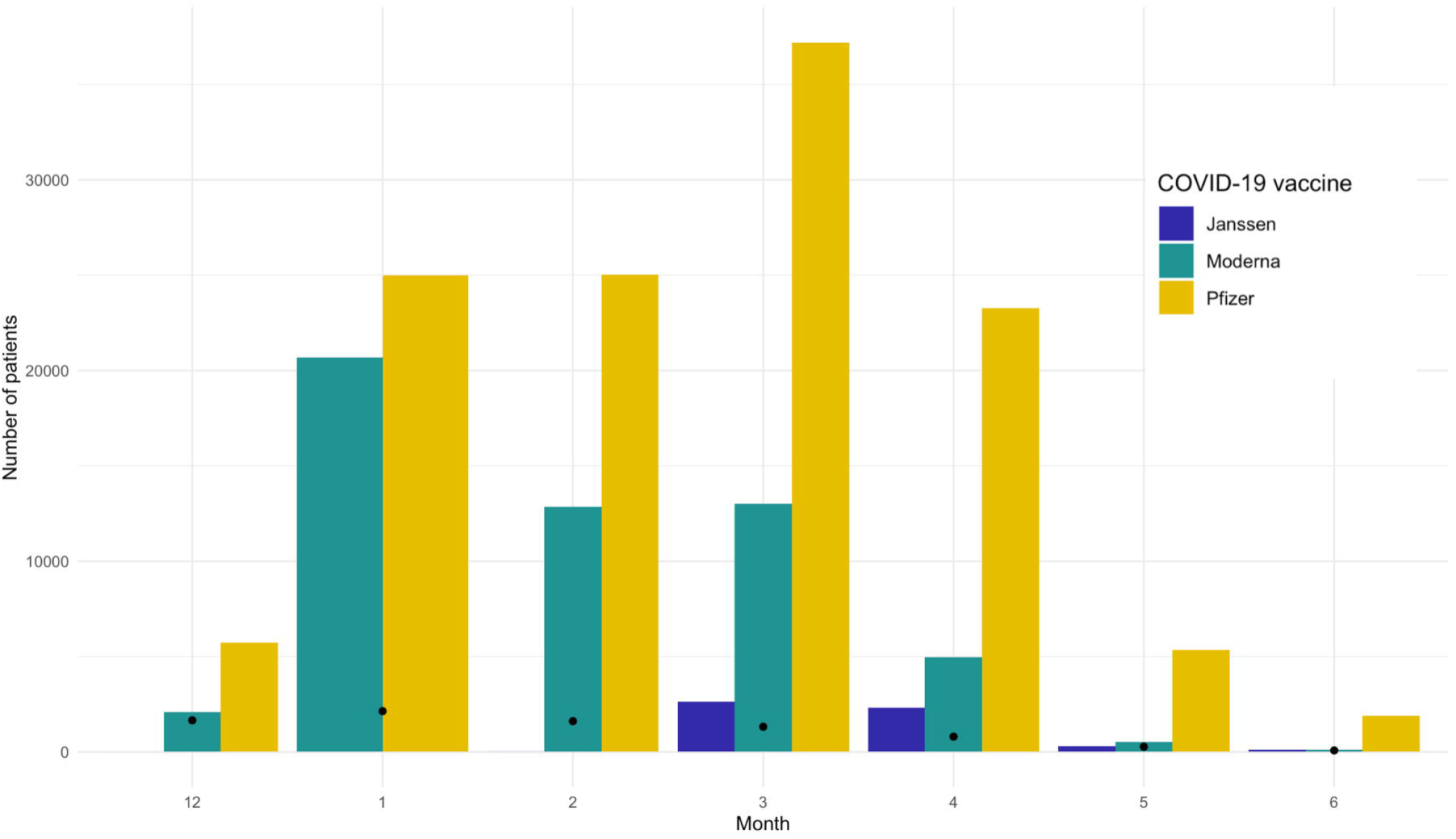
Diagnostics for the absolute effectiveness study comparing the cohort vaccinated with at least one dose of Pfizer, Moderna or Janssen COVID-19 vaccines and unvaccinated cohort anchored on a date or on a visit: (A) covariate balance before and after propensity score matching, (B) preference score balance and (C) effect of negative control calibration displaying effect estimate and standard error. In (A), each dot represents the standardized difference of the means for a single covariate before and after stratification on the propensity score. In (C), each blue dot is a negative control. The area below the dashed line indicates estimates with p<0.05 and the orange area indicates estimates with calibrated p<0.05.

Patients vaccinated with Pfizer-BioNTech had a similar distribution of baseline characteristics compared to the patients vaccinated with Moderna but differed from the patients vaccinated with Janssen. On average, the latter group was older, had more patients with race recorded as Black, and had more co-morbidities such as diabetes mellitus or hypertensive disorder (Table 1).

### Main week-by-week absolute effectiveness analysis

Figure 3, A shows week-by-week estimates for patients vaccinated with at least one dose of Pfizer-BioNTech or Moderna. Due to the small sample size, we were not able to obtain stable week-by-week estimates for Janssen. While week one was characterized by unexpectedly high effectiveness (58%, 95% CI 45-69% against COVID-19 infection and 72%, 95% CI 57-83% against COVID-19 associated hospitalization), we observed plausible increasing effectiveness beginning week 2 with the effectiveness on week 6 approximating 84% (95% CI 72-91%) for COVID-19 infection and 86% (95% CI 69-95) for COVID-19-associated hospitalization.

**Figure 3.**
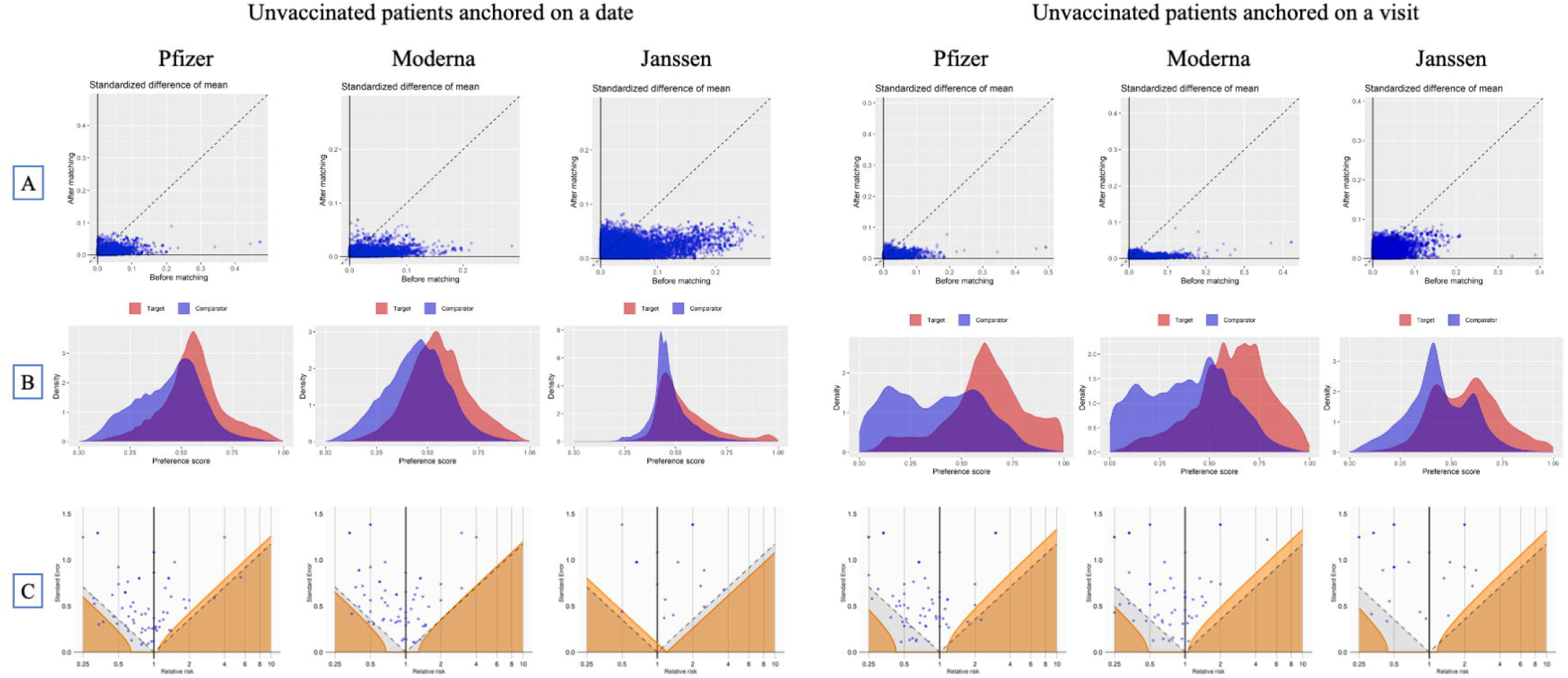
Week-by-week estimates of vaccine effectiveness of Pfizer-BioNTech and Moderna after 1^st^ dose, % and 95% CI for COVID-19 infection (A) and COVID-19 hospitalization (B). Chart review of COVID-19 cases (defined as a positive COVID-19 test) during week 1, vaccinated and unvaccinated patients (C).

We then looked at the week one COVID-19 infection cases to explain high effectiveness. A chart review of week1 positive COVID-19 tests revealed a high proportion of unvaccinated patients seeking care related to COVID-19 symptoms or COVID-19 exposure (85% in total) compared to only 69% of vaccinated patients. Initial healthcare encounters in vaccinated population were oftentimes related to other medical reasons such as co-morbid conditions or surgeries (39% compared to 21% in unvaccinated population, Appendix 5). Moreover, an observed gap between symptom onset and an initial healthcare encounter was more pronounced in the vaccinated cohort as the patients attributed their symptoms to temporal vaccine side effects as opposed to COVID-19 infection.

When looking at the severity of COVID-19 symptoms at the initial encounter during week one after the index date, we observed that the unvaccinated cohort had a higher proportion of asymptomatic cases (39% compared to 11%) while the vaccinated population had more severe or mild cases (34% and 48% respectively).

### Sensitivity analysis

#### Overall effectiveness

As cohort analysis allows us to construct Kaplan-Meier curves to assess effectiveness over time, we also looked at the effectiveness during the year after the first dose (Figure 4). We observed similar trends with all three vaccines being less effective during the first month after the first dose. After that, Pfizer-BioNTech and Moderna were highly effective against both COVID-19 infection and COVID-19 associated hospitalization, while Janssen vaccine exhibited a wide range of effectiveness (Appendix 6). The results for fully vaccinated patients with time-at-risk starting at the full vaccination matched the results of the clinical trials for corresponding vaccines (detailed estimates are provided in Appendix 7 and 8).

**Figure 4.**
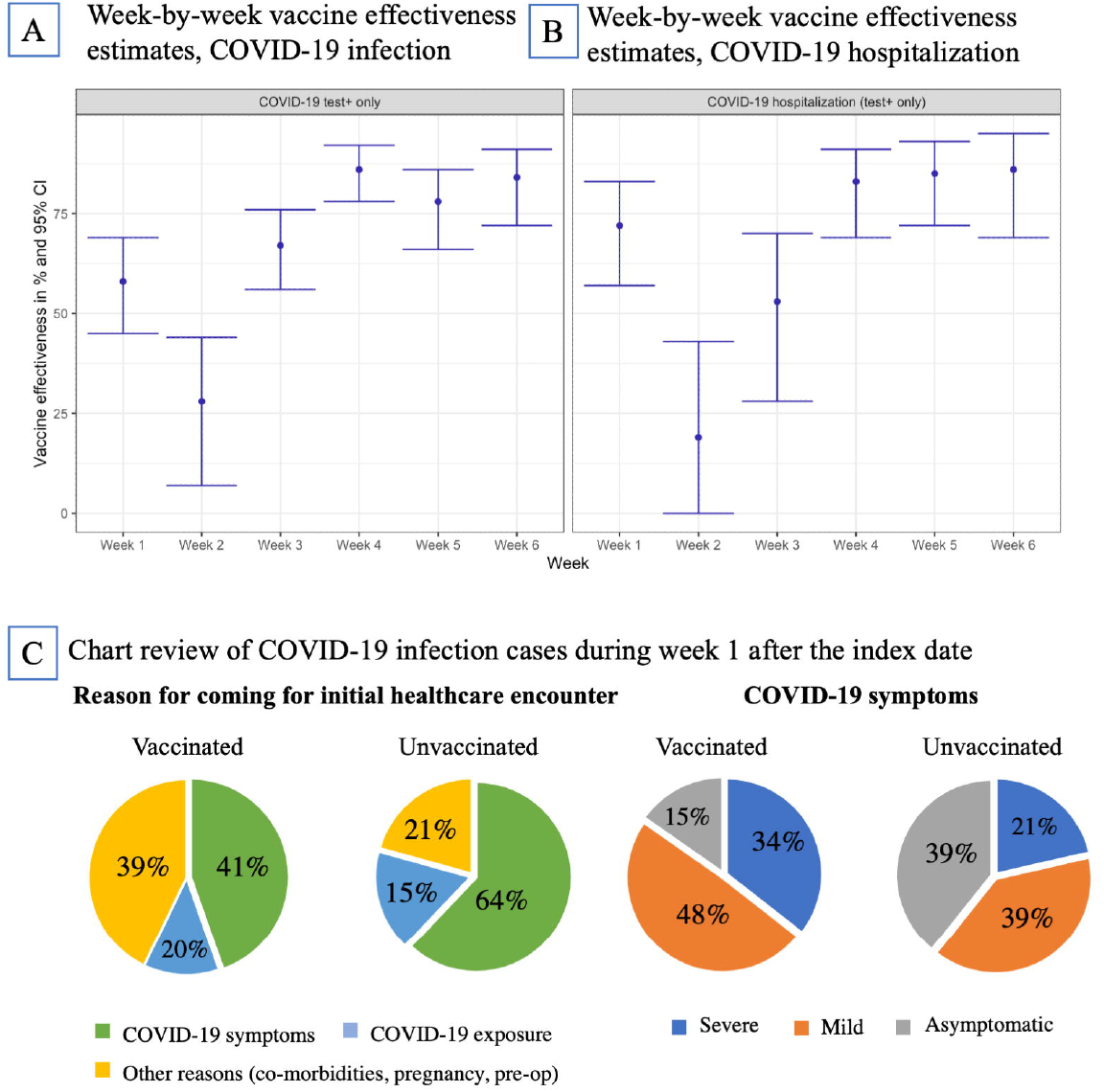
Kaplan-Meier curves for absolute effectiveness of COVID-19 vaccines for time-at-risk of 1 day – 365 days after the first dose compared to the unvaccinated patients residing in New York City.

Our initial design included a positive COVID-19 test or a diagnostic code as an outcome. Upon further case examination, we discovered that COVID-19 diagnostic codes in the CUIMC data were partially assigned to the patients with negative COVID-19 tests on or immediately following the date of diagnosis. In that case, ICD10CM code U07.1 “Disease caused by Severe acute respiratory syndrome coronavirus 2” was entered in the system for billing purposes (COVID-19 molecular or antibody tests) or for COVID-19 sequelae. We, therefore, focused on positive COVID-19 test only for our primary outcome, which led to higher effectiveness for all vaccines compared to using both positive test and diagnosis (Appendix 6).

Finally, exclusion of patients with prior COVID-19 infection in our main analysis resulted in higher absolute effectiveness. Inclusion of patients regardless of their prior COVID-19 status led to a small decrease in observed effectiveness (Appendix 9) for both COVID-19 infection and hospitalization in patients vaccinated with Moderna or Janssen.

## DISCUSSION

In this retrospective cohort study, we examined the weekly effectiveness of COVID-19 mRNA vaccines as well as the overall effectiveness of three COVID-19 vaccines commonly used in the US. COVID-19 mRNA vaccines were highly effective against both COVID-19 infection and COVID-19 associated hospitalization. Our findings support the RCTs and previously published post-marketing studies for all three vaccines. Larger sample size for patients vaccinated with COVID-19 mRNA vaccines allowed us to have more power, which resulted in overlapping yet narrower confidence intervals compared to the RCTs. On the other hand, our study had fewer patients with the Janssen vaccine, which resulted in wider yet overlapping intervals compared to the Janssen’s vaccine RCT ^1(p26),2,7^.

Our study complemented previous studies by examining and comparing disparate design choices such as studying both COVID-19-associated hospitalization and COVID-19 infection, different outcome definitions and broad age group ^4,5^. We scrutinized the effectiveness of the mRNA vaccines following the first dose and confirmed the findings of moderate vaccine effectiveness during the first two weeks. For week one following the first dose we discovered previously uncaptured differential biases in vaccinated and unvaccinated populations. Vaccination directly influenced the attitude of patients towards their symptoms, causing a delay in seeking care and a higher symptom severity threshold needed to seek care. Mild COVID-19 cases following vaccination were mainly captured upon seeking outpatient and inpatient care for other conditions. Such a difference may affect any observational vaccine study that uses hospitalization as a surrogate for COVID-19 severity.

Our sensitivity analyses discovered several challenges and potential biases that must be accounted for when conducting vaccine effectiveness studies on observational data. First, we observed that outcome definitions are prone to measurement error, which has not been studied thoroughly. The specifics of data capture and billing processes were associated with some patients having assigned COVID-19 diagnosis codes for billing for tests rather than as an indicator of active disease. Another reason for assigning the code was COVID-19 sequela, where the actual date of COVID-19 infection could have been anywhere from 6 months to a couple of weeks in the past. Such index date misclassification can be present in other healthcare institutions and therefore should be scrutinized to make valid inferences.

Second, inclusion or exclusion of patients with prior COVID-infection influenced estimated effectiveness. We observed that inclusion of patients with prior COVID-19 leads to lower effectiveness for all vaccines regardless of the outcome definition.

If absolute effectiveness is studied, an appropriate index event (anchor) for the unvaccinated cohort must be chosen. In our study, we observed that a date represents a better counterfactual than a medical visit for COVID-19 vaccination which is reflected in propensity score balance and covariate balance. Nevertheless, other institutions may have different vaccination pathways such as vaccination on discharge, which can make a visit a better counterfactual for vaccination. More generally, completeness of vaccination data capture is a crucial feature that influences the robustness of the study. While CUIMC data ensures complete exposure capture by linking EHR to the City and State Registries, the researchers should exhibit caution with conducting studies on the data sources with unknown vaccination capture.

We obtained similar results to RCTs, which strengthens the conclusions about the high effectiveness of vaccines against COVID-19 infection in the broad age group. While these RCTs allowed us to make such comparisons for absolute effectiveness, there are other research questions for which RCTs may not be feasible or readily available. The US and international booster campaigns raise the question of vaccine comparative effectiveness to prioritize vaccination. An indirect comparison may not be accurate due to the differences in the populations we observed in our study. First, patients vaccinated with Janssen were substantially different from mRNA patients: on average, they were older, had a higher proportion of patients with race recorded as Black and had more comorbidities. Therefore, comparative effectiveness studies of Janssen and mRNA vaccines require robust techniques such as large-scale propensity matching to ensure valid comparison. Second, while Modena and Pfizer patients had similar baseline characteristics, the temporal distribution of vaccinations in CUIMC data differ. Moderna vaccine was administered early on in 2021 with the peak in January, while Pfizer vaccination peaked in April. Given the varying baseline COVID-19 prevalence, a comparison of mRNA vaccines requires matching patients on calendar month to account for this potential bias. These vaccines also had different administration pathways in our system. As opposed to Pfizer vaccine, which was administered at Columbia University Irving Medical Center/New York-Presbyterian sites to all patients over a prolonged period, Moderna vaccination was performed elsewhere and recorded for actively observed patients. Such patients were more likely to get tested or receive care outside of our healthcare system.

## LIMITATIONS

Due to observational nature of the study, the data sources may not have complete capture of patient conditions, which was mitigated by having free and available COVID-19 testing in Columbia University Irving Medical Center/New York-Presbyterian sites as well as by having data capture from New York City and State Immunization Registries. Along with availability of testing, COVID-19 baseline infection rate difference was mitigated by matching the target and comparator groups on the index date and using the index month as a covariate in propensity score model. While our outcome phenotype algorithms may be subject to measurement error, we provided additional sensitivity analyses with alternative outcome definitions.

## CONCLUSIONS

Observational data can be used to ascertain vaccine effectiveness if potential biases such as exposure and outcome misclassification are accounted for, and appropriate anchoring event is selected. When analyzing granular vaccine effectiveness researchers need to scrutinize the data to ensure that compared groups exhibit similar health seeking behavior and are equally likely to be captured in the data. Given the difference in temporal trends of vaccine exposure and baseline characteristics, there is a need for large-scale direct comparison of vaccines to examine comparative effectiveness.

## Data Availability

All data produced in the present work are contained in the manuscript

## DECLARATION

## Author contribution

GH designed and supervised the study. All co-authors contributed to interpretation of the results and writing the manuscript, approved the final version and had final responsibility for the decision to submit for publication.

## Acknowledgment

We would like to acknowledge Patrick Ryan, an employee of Janssen Research and Development, Titusville, New Jersey, for his thoughtful feedback on the study.

## Ethical approval

The protocol for this research was approved by the Columbia University Institutional Review Board (AAAO7805).

