## Appendix for "COVID-19 vaccination effectiveness rates by week and sources of bias"

**Supplementary materials**

**Appendix 1.** Data source description

The Columbia University Irving Medical Center (CUIMC) database comprises electronic health records on more than 6 million patients, with data collection starting in 1985. CUIMC is a Northeast US quaternary care center with primary care practices in northern Manhattan and surrounding areas, and the database includes inpatient and outpatient care. The database currently holds information about the person (demographics), visits (inpatient and outpatient), conditions (billing diagnoses and problem lists), drugs (outpatient prescriptions and inpatient orders and administrations), devices, measurements (laboratory tests and vital signs), and other observations (symptoms). The data sources include current and previous electronic health record systems (homegrown Clinical Information System, homegrown WebCIS, Allscripts Sunrise Clinical Manager, Allscripts TouchWorks, Epic Systems), administrative systems (IBM PCS-ADS, Eagle Registration, IDX Systems, Epic Systems), and ancillary systems (homegrown LIS, Sunquest, Cerner Laboratory). Additionally, it contains the information on vaccination from New York City and State immunization registries.

**Appendix 2.** Retrospective cohort COVID-19 vaccine effectiveness study design overview.

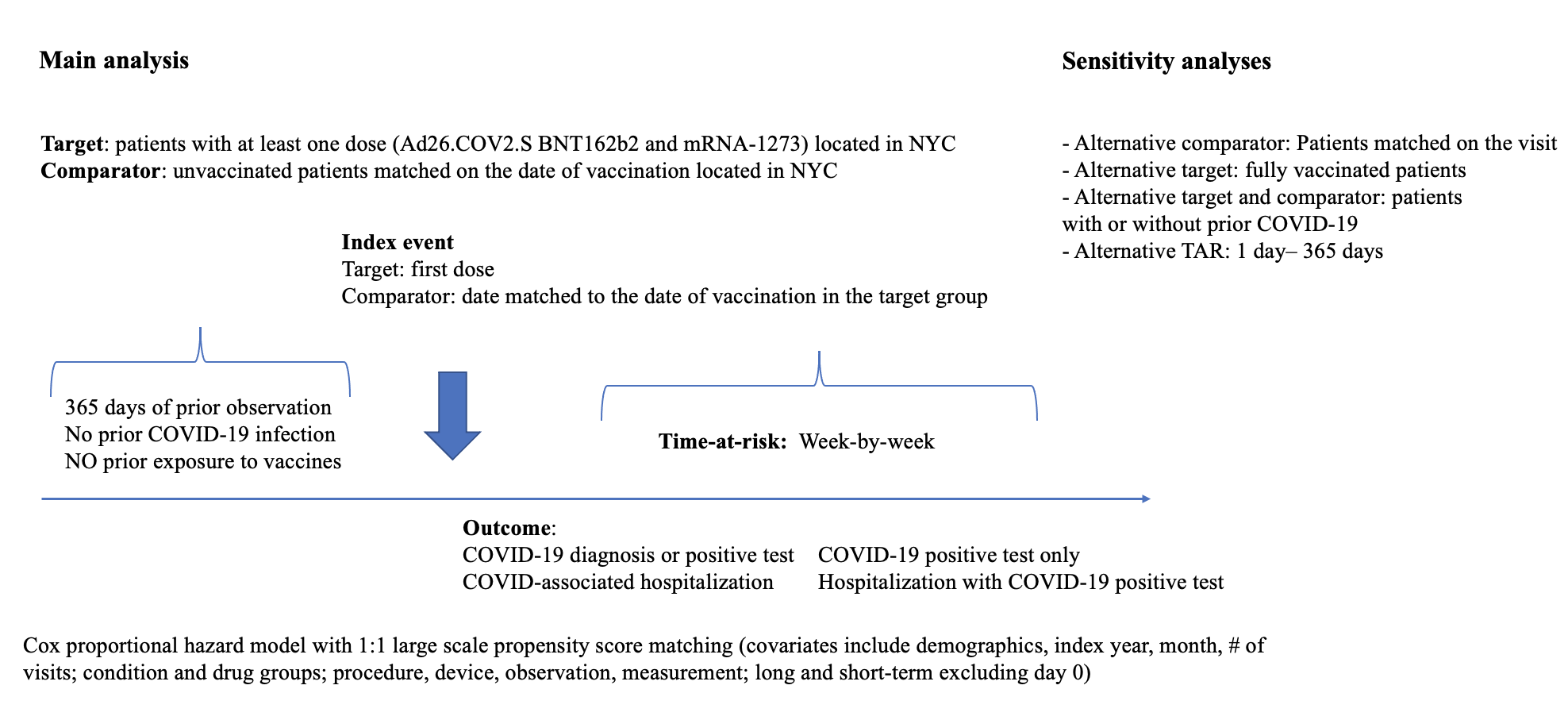

**Appendix 3.** Cohort definitions and codes for the absolute COVID-19 vaccine effectiveness study­­

**3.1** Cohort definitions for target comparator and outcome cohorts for studying absolute effectiveness of COVID-19 vaccines.

|  | **Definition and link to the public repository** |
| --- | --- |
| Target cohorts | Target cohorts were defined as patients with at least one dose of the corresponding vaccine (Pfizer, Moderna, Janssen)  Index event: first exposure to the corresponding vaccine  Inclusion and exclusion criteria:  - 365 days of prior observation  - no other COVID-19 vaccine exposure in 120 days prior and 120 days after the index date  - no prior COVID-19 infection (diagnosis code of COVID-19 or positive test)  - residence in New York City determined by the zip code recorded  For the analysis on fully vaccinated patients, we applied the same criteria and required patients to have a) the second dose of Pfizer or Moderna vaccine (if applicable) within 14 to 56 days after the first dose b) at least 14 days of observation after the second dose (one dose of Janssen).  **Links:**  <https://atlas.ohdsi.org/#/cohortdefinition/498>  <https://atlas.ohdsi.org/#/cohortdefinition/494>  <https://atlas.ohdsi.org/#/cohortdefinition/497>  <https://atlas.ohdsi.org/#/cohortdefinition/418>  <https://atlas.ohdsi.org/#/cohortdefinition/417>  <https://atlas.ohdsi.org/#/cohortdefinition/420> |
| Comparator cohorts | Comparator cohorts were created separately for each target cohort by selecting patients with no COVID-19 vaccination in their record (any vaccine), 365 days of prior observation and New York City residence. The patients were matched on the index date of one of the target group participants for the comparator anchored on a date and on the date of a healthcare encounter within 3-day corridor for the comparator anchored on a visit. |
| Outcome cohorts | For the main analysis COVID-19 infection was defined as a COVID-19 test with the result ‘Positive’ or ‘Detected’.  COVID-19 associated hospitalization was defined as an inpatient, emergency department or intensive care unit admission with a positive COVID-19 test recorded within 30 days prior or during hospitalization.  For a sensitivity analysis we applied the abovementioned criteria with adding COVID-19 diagnosis as an alternative for positive COVID-19 test.  Links:  <https://atlas.ohdsi.org/#/cohortdefinition/425>  <https://atlas.ohdsi.org/#/cohortdefinition/422> |

**3.2** Codes used in the study.

1. **Pfizer vaccine:**

RxNorm 2468235	SARS-CoV-2 (COVID-19) vaccine, mRNA-BNT162b2 0.1 MG/ML Injectable Suspension

2. **Moderna vaccine:**

RxNorm 2470234	SARS-CoV-2 (COVID-19) vaccine, mRNA-1273 0.2 MG/ML Injectable Suspension

3. **Janssen vaccine:**

CVX 212	SARS-COV-2 (COVID-19) vaccine, vector non-replicating, recombinant spike protein-Ad26, preservative free, 0.5 mL

4. **COVID-19 diagnosis:**

ICD10-CM U07.1 Emergency use of U07.1 | COVID-19

5. **COVID-19 test:**

LOINC 94500-6 SARS-CoV-2 (COVID-19) RNA [Presence] in Respiratory specimen by NAA with probe detection

LOINC 94558-4 SARS-CoV-2 (COVID-19) Ag [Presence] in Respiratory specimen by Rapid immunoassay

**Appendix 4.** Negative controls

| **SNOMED concept id** | **SNOMED concept name** |
| --- | --- |
| 438945 | Accidental poisoning by benzodiazepine-based tranquilizer |
| 434455 | Acquired claw toes |
| 316211 | Acquired spondylolisthesis |
| 201612 | Alcoholic liver damage |
| 438730 | Alkalosis |
| 441258 | Anemia in neoplastic disease |
| 432513 | Animal bite wound |
| 4171556 | Ankle ulcer |
| 4098292 | Antiphospholipid syndrome |
| 77650 | Aseptic necrosis of bone |
| 4239873 | Benign neoplasm of ciliary body |
| 23731 | Benign neoplasm of larynx |
| 199764 | Benign neoplasm of ovary |
| 195500 | Benign neoplasm of uterus |
| 4145627 | Biliary calculus |
| 4108471 | Burn of digit of hand |
| 75121 | Burn of lower leg |
| 4284982 | Calculus of bile duct without obstruction |
| 434327 | Cannabis abuse |
| 78497 | Cellulitis and abscess of toe |
| 4001454 | Cervical spine ankylosis |
| 4068241 | Chronic instability of knee |
| 195596 | Chronic pancreatitis |
| 4206338 | Chronic salpingitis |
| 4058397 | Claustrophobia |
| 74816 | Contusion of toe |
| 73302 | Curvature of spine |
| 4151134 | Cyst of pancreas |
| 77638 | Displacement of intervertebral disc without myelopathy |
| 195864 | Diverticulum of bladder |
| 201346 | Edema of penis |
| 200461 | Endometriosis of uterus |
| 377877 | Esotropia |
| 193530 | Follicular cyst of ovary |
| 4094822 | Foreign body in respiratory tract |
| 443421 | Gallbladder and bile duct calculi |
| 4299408 | Gouty tophus |
| 135215 | Hashimoto thyroiditis |
| 442190 | Hemorrhage of colon |
| 43020475 | High risk heterosexual behavior |
| 194149 | Hirschsprung's disease |
| 443204 | Human ehrlichiosis |
| 4226238 | Hyperosmolar coma due to diabetes mellitus |
| 4032787 | Hyperosmolarity |
| 197032 | Hyperplasia of prostate |
| 140362 | Hypoparathyroidism |
| 435371 | Hypothermia |
| 138690 | Infestation by Pediculus |
| 4152376 | Intentional self poisoning |
| 192953 | Intestinal adhesions with obstruction |
| 196347 | Intestinal parasitism |
| 137977 | Jaundice |
| 317510 | Leukemia |
| 765053 | Lump in right breast |
| 378165 | Nystagmus |
| 434085 | Obstruction of duodenum |
| 4147016 | Open wound of buttock |
| 4129404 | Open wound of upper arm |
| 438120 | Opioid dependence |
| 75924 | Osteodystrophy |
| 432594 | Osteomalacia |
| 30365 | Panhypopituitarism |
| 4108371 | Peripheral gangrene |
| 440367 | Plasmacytosis |
| 439233 | Poisoning by antidiabetic agent |
| 442149 | Poisoning by bee sting |
| 4314086 | Poisoning due to sting of ant |
| 4147660 | Postural kyphosis |
| 434319 | Premature ejaculation |
| 199754 | Primary malignant neoplasm of pancreas |
| 4311499 | Primary malignant neoplasm of respiratory tract |
| 436635 | Primary malignant neoplasm of sigmoid colon |
| 196044 | Primary malignant neoplasm of stomach |
| 433716 | Primary malignant neoplasm of testis |
| 133424 | Primary malignant neoplasm of thyroid gland |
| 194997 | Prostatitis |
| 80286 | Prosthetic joint loosening |
| 443274 | Psychostimulant dependence |
| 314962 | Raynaud's disease |
| 37018294 | Residual osteitis |
| 4288241 | Salmonella enterica subspecies arizonae infection |
| 45757269 | Sclerosing mesenteritis |
| 74722 | Secondary localized osteoarthrosis of pelvic region |
| 200348 | Secondary malignant neoplasm of large intestine |
| 43020446 | Sedative withdrawal |
| 74194 | Sprain of spinal ligament |
| 4194207 | Tailor's bunion |
| 193521 | Tropical sprue |
| 40482801 | Type II diabetes mellitus uncontrolled |
| 74719 | Ulcer of foot |
| 196625 | Viral hepatitis A without hepatic coma |
| 197494 | Viral hepatitis C |
| 4284533 | Vitamin D-dependent rickets |

Link to the original list of negative controls used in EUMAEUS study: <https://ohdsi-studies.github.io/Eumaeus/Protocol.html#8_Research_Methods>

**Appendix 5.** Summary of manual chart review of COVID-19 infection cases during week 1 after the index date, patients vaccinated with mRNA vaccines and unvaccinated patients.

|  | **Pfizer- BioNTech** | **Moderna** | **Pfizer- BioNTech and Moderna** | **Unvaccinated**  **patients** |
| --- | --- | --- | --- | --- |
| Total | 36 | 25 | 61 | 28 |
| Average age | 65 | 67.8 | 65.8 | 58 |
| ***COVID-19 symptoms*** | | | | |
| Severe | 14 (39%) | 7 (28%) | 21 (34%) | 6 (21%) |
| Mild | 18 (50%) | 11 (44%) | 29 (48%) | 11 (39%) |
| Asymptomatic | 2 (6%) | 7 (28%) | 9 (15%) | 11 (39%) |
| ***Reason for coming for initial healthcare encounter*** | | | | |
| COVID-19 symptoms | 17 (47%) | 8 (32%) | 25 (41%) | 18 (64%) |
| Exposure to COVID-19 | 3 (8%) | 4 (16%) | 7 (11%) | 5 (18%) |
| For other reason (co-morbidities, procedures etc.) | 13 (36%) | 11 (44%) | 24 (39%) | 6 (21%) |
| ***Type of initial healthcare encounter*** | | | | |
| Telehealth/phone | 5 (14%) | 6 (24%) | 11 (18%) | 3 (11%) |
| Test only | 3 (8%) | 2 (8%) | 5 (8%) | 6 (21%) |
| OP | 4 (11%) | 3 (12%) | 7 (11%) | 1 (4%) |
| ED or IP | 24 (67%) | 14 (56%) | 38 (62%) | 18 (64%) |

**Appendix 6.** Estimates for absolute effectiveness of COVID-19 vaccines for time-at-risk of 1 day – 365 days after the first dose in the vaccinated patients without prior COVID-19 infection compared to unvaccinated patients residing in NYC.

|  | **COVID-19 infection** | | **COVID-19 hospitalization** | | **COVID-19 positive test only** | | **COVID-19 positive test only hospitalization** | |
| --- | --- | --- | --- | --- | --- | --- | --- | --- |
|  | VE (95% CI), % | P-value | VE (95% CI), % | P-value | VE (95% CI), % | P-value | VE (95% CI), % | P-value |
| **Pfizer-** **BioNTech** | 42 (37 – 47) | <0.01 | 63 (56-70) | <0.01 | 71 (66 - 75) | <0.01 | 69 (62 - 75) | <0.01 |
| **Moderna** | 54 (48 – 60) | <0.01 | 76 (69 – 82) | <0.01 | 78 (73 – 83) | <0.01 | 81 (74 – 87) | <0.01 |
| **Janssen** | 24 (0-55) | 0.31 | 64 (0.1 – 1.06) | 0.09 | 53 (0 – 82) | 0.1 | 70 (2 – 93) | 0.08 |

**Appendix 7.** Estimates for absolute effectiveness of COVID-19 vaccines for time-at-risk of 1 day – 365 days after full vaccination in fully vaccinated patients without prior COVID-19 infection compared to unvaccinated patients residing in NYC.

|  | **COVID-19**  **positive test only** | | **COVID-19 positive test only hospitalization** | | **COVID-19 infection** | | **COVID-19 hospitalization** | |
| --- | --- | --- | --- | --- | --- | --- | --- | --- |
|  | VE (95% CI), % | P-value | VE (95% CI), % | P-value | VE (95% CI), % | P-value | VE (95% CI), % | P-value |
| **Pfizer-** **BioNTech** | 94 (91- 95) | <0.01 | 95 (92-97) | <0.01 | 70 (66-74) | <0.01 | 88 (84-92) | <0.01 |
| **Moderna** | 97 (94-98) | <0.01 | 96 (92-99) | <0.01 | 72 (66 – 77) | <0.01 | 92 (87-95) | <0.01 |
| **Janssen** | 81 (50-94) | <0.01 | 92 (58-100) | 0.03 | 55 (23 – 75) | 0.01 | 87 (56-98) | 0.01 |

**Appendix 8.** Comparison of the absolute effectiveness estimates in fully vaccinated patients obtained in our study and those from the randomized clinical trials of the corresponding vaccines.

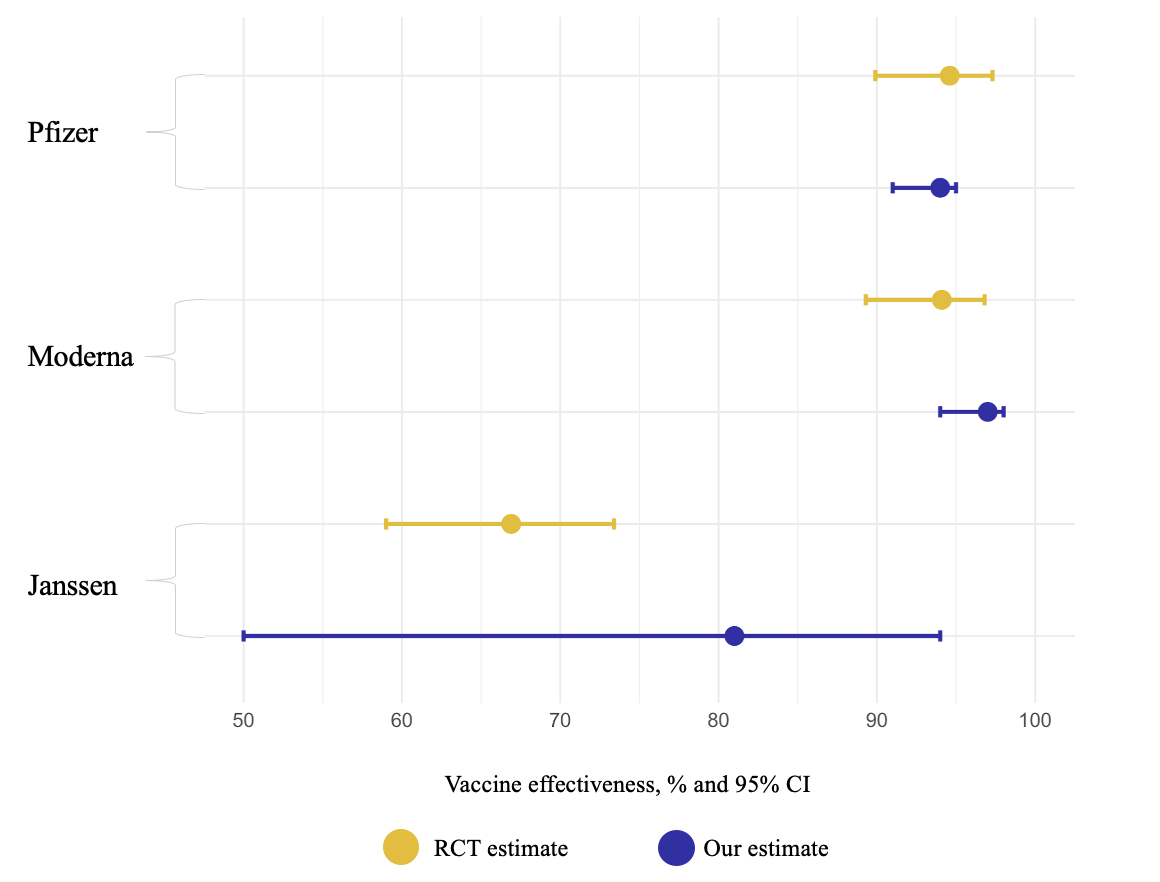

**Appendix 9.** Estimates for absolute effectiveness of COVID-19 vaccines for time-at-risk of 1 day – 365 days after the first dose in the vaccinated patients with or without prior COVID-19 infection compared to unvaccinated patients residing in NYC.

|  | **COVID-19 infection** | | **COVID-19 hospitalization** | | **COVID-19 positive test only** | | **COVID-19 positive test only hospitalization** | |
| --- | --- | --- | --- | --- | --- | --- | --- | --- |
|  | VE (95% CI), % | P-value | VE (95% CI), % | P-value | VE (95% CI), % | P-value | VE (95% CI), % | P-value |
| **Pfizer-** **BioNTech** | 43 (38-48) | <0.01 | 64 (57-70) | <0.01 | 71 (66-75) | <0.01 | 71(64-76) | <0.01 |
| **Moderna** | 51 (45-57) | <0.01 | 71 (63-78) | <0.01 | 76 (71-81) | <0.01 | 81 (73-86) | <0.01 |
| **Janssen** | 15 (0-49) | 0.52 | 60 (2-86) | 0.06 | 45 (0-75) | 0.12 | 63 (0-90) | 0.09 |
